# When Do Drug Shortages Raise Acquisition Costs? Average Duration Effects and Heterogeneous Price Pass-Through

**DOI:** 10.64898/2026.07.27.26359030

**Authors:** Qiaoyuan Li

## Abstract

Drug shortages represent persistent supply disruptions in the U.S. pharmaceutical market, threatening patient access and increasing drug costs. Prior research commonly treats shortages as binary events and relies on static designs, limiting insight into how shortage characteristics drive cost escalation. This study uncovers the heterogeneity behind drug shortages and pharmacy acquisition costs of generic non-injectable drugs. FDA drug shortage records with weekly National Average Drug Acquisition Cost (NADAC) prices were fit with fixed-effects models, duration-specific models, and a double machine-learning framework to characterize heterogeneity in shortage-price associations by duration, market structure, and shortage reasons. In the baseline two-way fixed-effects model, active shortage designation alone was not associated with a significant increase in NADAC under clustered standard errors. In duration-specific models, shortages lasting more than four consecutive weeks were associated with approximately 7% higher NADAC, while each additional cumulative shortage week was associated with approximately 0.37% higher NADAC. Estimated CATEs varied widely across drugs in each week. Allocation restrictions, raw material and distribution disruptions, together with a lack of manufacturers, characterized shortages with higher estimated CATEs. These findings support monitoring both shortage persistence and supply-chain mechanisms to mitigate impacts on healthcare systems.

## 1. Introduction

Drug shortages remain a persistent challenge in the U.S. pharmaceutical supply chain, disrupting access to essential medications for retail pharmacies and health systems and placing strain on managed care budgets, pharmacy benefit managers, and patients. When shortages occur, pharmacies and hospitals face constrained sourcing options and may be forced to activate higher-cost secondary suppliers, increasing downstream costs for payers [6]. These pressures have intensified in recent years: the COVID-19 pandemic exposed deep vulnerabilities in pharmaceutical production and distribution networks, and active shortage counts reached decade-high levels by 2022–2023 [29, 32]. Despite their growing frequency and economic consequence, the conditions under which shortages are associated with meaningful price escalation remain poorly understood.

One critical challenge in this literature is confounding. Drugs that experience shortages are structurally different from those that do not, and they tend to be older generic products manufactured by fewer suppliers, often operating in markets with thin margins and limited redundancy. Naive cross-sectional comparisons of shortage versus non-shortage drugs therefore capture structural price differences that predate and persist independently of the shortage event. Even when price increases coincide with shortages, it is difficult to distinguish between (a) price increases associated with the supply disruption and (b) pre-existing pricing differences or common shocks that coincide with shortage designation.

A second challenge is heterogeneity. Shortages vary widely in duration. Some resolve within weeks, others persist for months or years. A manufacturing delay with a clear recovery timeline creates different market dynamics than a shortage driven by active rationing, where manufacturers allocate scarce supply across customers under quota-like arrangements. If these different shortage types carry fundamentally different price implications, then average effect estimates will be misleading both for policy and for managed care monitoring.

This paper addresses both challenges. To improve within-drug comparability, we adopt a two-way fixed-effects design: comparing each drug to itself over time, restricting to drugs that transition between shortage and non-shortage states during the study period. This eliminates time-invariant drug heterogeneity as a source of confounding. To capture heterogeneity, we deploy a Causal Forest Double Machine Learning estimator with three levels of features, including market structure, FDA rule-based text flags, and ML-estimated text mechanism scores. These features help us estimate drug-week variation in the relationship between shortage and price.

We find that drug shortages are not uniformly associated with increases in pharmacy acquisition costs. Average associations are concentrated among prolonged shortages and are modest for short disruptions: shortages lasting more than four consecutive weeks were associated with approximately 7% higher NADAC. But the distribution of estimated drug-week price responses is wide, spanning roughly 40 percentage points between the bottom and top deciles. This heterogeneity is driven primarily by allocation and supply-chain constraint language in FDA notifications, not by duration alone. The highest estimated price responses are typically observed in shorter shortage periods characterized by active supply rationing. These findings suggest that managed care monitoring and policy responses are encouraged to track both shortage persistence and FDA-reported mechanism signals.

## 2. Literature Review

### 2.1 Drug Shortages and Market Dynamics

Drug shortages in the United States have increased substantially since the 2000s, with generic sterile injectables disproportionately affected due to the high capital costs of sterile manufacturing, narrow profit margins, and limited supplier redundancy [1, 2, 36]. The market structure of generic pharmaceuticals creates inherent instability: when one manufacturer encounters a production problem, the capacity to substitute is often constrained by the small number of remaining suppliers, and by the time required to scale production or obtain regulatory approval for alternative sources [3, 22, 23]. Generic markets characterized by thin margins, limited manufacturing redundancy, and concentrated supplier structures are especially exposed to disruptions [22, 23, 30, 33]. Competition in the global supply of active pharmaceutical ingredients remains concentrated, with a substantial share of shortage-prone generics reliant on a small number of foreign manufacturing facilities [30, 33].

The FDA Drug Shortages database provides a publicly available record of nationally recognized supply disruptions [13], including product information, shortage onset and resolution dates, and free-text narratives explaining the disruption mechanism. Prior research has documented associations between shortage incidence and price changes, but study designs have varied widely in their ability to control for confounding [4, 5, 19]. National reviews and managed care commentary likewise underscore the persistence of U.S. shortages and their relevance for pharmacy and drug spending [20, 21, 31]. Comparative analyses confirm that U.S. shortages are more persistent and more frequently driven by manufacturing quality deficiencies than those observed in Canada and other high-income countries [26, 27]. Cross-sectional studies that compare shortage to non-shortage drugs at a point in time often fail to account for the structural differences between these groups. Before-after studies that track individual drug prices around shortage events are closer to within-drug identification but may still be contaminated by time trends or coincident market changes [17].

### 2.2 Price Effects of Drug Shortages

Hernandez et al. [4] documented significant price increases in drugs experiencing shortages in a retrospective study, finding that shortage drugs had higher prices than non-shortage comparators. Dave et al. [5] found that market concentration in generic drug markets, a structural driver of shortage vulnerability, was associated with higher prices, linking the conditions that make shortages likely to the conditions that make price escalation possible. Conti and Wosinska [23] emphasize that thin margins and limited competition in generic markets constrain manufacturers’ ability to absorb production disruptions without exiting supply. Mulcahy et al. [6] estimated that drug shortages impose substantial cost increases on healthcare consumers. International evidence likewise documents shortage-related price pressures, though institutional settings differ from U.S. retail pharmacy markets [37]. However, these studies generally estimate average associations and do not decompose effects by shortage type, duration, or mechanism.

The role of shortage duration has received limited systematic investigation. Economic intuition suggests that short disruptions may be absorbed through inventory buffers, formulary switching, or therapeutic substitution strategies available to health systems and pharmacy benefit managers [27, 34], while prolonged disruptions exhaust these strategies and reduce effective competition [22]. Policy analyses have linked reimbursement incentives and supply-chain fragility to shortage persistence [38], reinforcing the importance of studying acquisition costs rather than list prices alone [14, 35]. Prior work has not cleanly distinguished the average cost implications of short versus prolonged shortages within a within-drug fixed-effects framework.

### 2.3. Machine Learning and Text-Based Methods in Health Economics

The application of machine learning methods to causal inference in health economics has grown rapidly. Double machine learning [8] provides a framework for nuisance function estimation that achieves valid inference under high-dimensional controls. The Causal Forest [9, 10] extends this framework to heterogeneous treatment effect estimation, allowing personalized treatment effects to vary nonparametrically across a feature space. These methods have been applied across a range of health policy settings but have seen limited application to pharmaceutical pricing and supply chain questions.

Natural language processing (NLP) methods have been applied to FDA adverse event reports, clinical notes, and regulatory filings for pharmacovigilance and policy analysis [11, 12]. The embedding of FDA shortage reason narratives as structured heterogeneity drivers transforms free-text mechanism descriptions into econometrically interpretable features, a methodological contribution of this study. Rule-based parsing, ML text classification, and keyword extraction are all deployed here to create a tiered feature set with varying levels of granularity and statistical interpretability.

### 2.4. Contribution of This Study

This study makes three contributions. First, it applies a within-drug two-way fixed-effects design to estimate average shortage-price associations while controlling for drug-level and time-level confounding, and documents duration dependence in those associations. Second, it deploys Causal Forest DML with cross-fitting to characterize heterogeneity in adjusted shortage-price associations across drug-weeks. Third, it introduces FDA shortage reason narratives as interpretable heterogeneity drivers, demonstrating that allocation and supply-chain constraint language provide predictive signal beyond duration alone. These new methods uncover new insights into how heterogeneous cost escalations are passed through from shortage events.

## 3. Data

### 3.1 FDA Drug Shortages Database

Drug shortage data were obtained from the U.S. Food and Drug Administration (FDA) Drug Shortages database [13], which reports nationally recognized shortage events at the drug product level. Each record includes the drug name, dosage form, shortage start date, resolution date (if applicable), and a free-text narrative field describing the reported reason for the disruption.

Shortage narratives cover a range of mechanisms including raw material supply problems, manufacturing disruptions, allocation to customers under rationing arrangements, distribution failures, and voluntary product discontinuation. Shortage records were matched to NADAC data at the National Drug Code (NDC) level using product name and NDC cross-references.

### 3.2 National Average Drug Acquisition Cost (NADAC)

NADAC data were obtained from the Centers for Medicare & Medicaid Services (CMS) [14], which publishes weekly invoice-level pharmacy acquisition costs at the NDC level based on pharmacy survey data. NADAC reflects actual post-discount prices paid by retail pharmacies, making it the most accurate available measure of pharmacy acquisition cost. Unlike Average Wholesale Price (AWP) or Wholesale Acquisition Cost (WAC), which are list prices with limited relationship to actual transaction prices, NADAC reflects the prices that pharmacies and ultimately payers and health plans effectively face when procuring drugs. The weekly frequency and NDC-level granularity of NADAC allow us to track high-frequency price dynamics in response to shortage events, including within-shortage dynamics as episodes evolve.

### 3.3 Sample Construction and Attrition

The analysis proceeded in three stages of sample construction, as summarized in Table 1. The full linked panel comprises 989,799 weekly observations across 38,291 NDCs from 2023 to 2025. Shortage exposure is rare in the full panel (approximately 0.4% of drug-weeks), reflecting that FDA-listed shortages affect a small fraction of all marketed products at any given time.

**Table 1.** Analytic Sample Attrition.

| Stage | NDCs | Drug-Weeks | Criterion |
| --- | --- | --- | --- |
| 1. Full panel | 38,291 | 989,799 | All NADAC–NDC weekly obs. linked to FDA shortage data (2023–2025) |
| 2. At-risk drugs | 245 | 5,633 | NDCs with $\geq 1$ observed shortage episode; enables within-drug comparison |
| 3. Generic trimmed (primary analytic sample) | 199 | 5,003 | Generic NDCs; overlap-trimmed by random forest propensity score; 61 calendar periods |

Estimating shortage effects in the full panel requires a credible counterfactual: the price a shortage drug would have had, absent its shortage. With so few shortage observations relative to the overall market, cross-product comparisons carry high risk of confounding.

To address this, we restrict to at-risk drugs, which are NDCs with at least one observed shortage period during the study period. This yields 245 NDCs and 5,633 drug-weeks, each product now contributing both shortage and non-shortage weeks for within-drug comparison. We further apply random-forest propensity score trimming to enforce common support between treated (shortage) and control (non-shortage) weeks within the at-risk generic product sample [18, 24], yielding the primary analytic sample of 199 NDCs, 5,003 drug-weeks, and 61 calendar periods. This staged restriction improves within-product comparability and common support rather than maximizing sample size.

## 4. Research Methodology

### 4.1 Estimation Framework

The central analytical challenge is that shortage-prone drugs differ structurally from non-shortage drugs in ways that are difficult to observe and control for fully. To address this, we adopt a two-way fixed-effects (FE) design that exploits within-drug variation over time. Let *Y_it_* denote the log NADAC per unit for drug *i* in week *t*, and let *D_it_* be an indicator equal to one if drug *i* is in active shortage in week *t*. The baseline specification is:

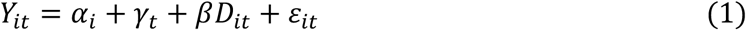

where *α_i_* is a drug (entity) fixed effect absorbing all time-invariant drug characteristics (generic status, therapeutic class, manufacturer concentration, formulation), and *γ_t_* is a calendar-week fixed effect absorbing all common time trends and aggregate shocks affecting all drugs simultaneously (e.g., supply chain disruptions, inflation, regulatory changes). The coefficient *β* summarizes the adjusted within-drug association between shortage status and log NADAC after removing time-invariant product differences and common weekly shocks. Standard errors are clustered at the NDC level to account for serial correlation within drug price series [25].

NADAC is not observed for every NDC in every calendar week; the panel therefore contains irregular observation timing that the FE framework accommodates through the available drug-week pairs. Two-way fixed-effects estimators can yield heterogeneous treatment-effect summaries when treatment timing varies across units [17].

### 4.2 Duration-Specific Effects

The baseline specification restricts shortage effects to be homogeneous across episodes regardless of duration. We augment the FE model to include four complementary duration and exposure measures:

1. active_shortage, the current-week shortage indicator;
2. current_shortage_duration, measuring consecutive weeks in the current uninterrupted spell;
3. cumulative_shortage_weeks, measuring total shortage weeks accumulated up to that point across all spells; and
4. long_shortage, a binary indicator equal to one when the current spell has lasted more than four consecutive weeks, a threshold reflecting the approximate horizon at which pharmacy inventory buffers and formulary switching are commonly exhausted.

Published shortage registries report median durations exceeding one year for oral generics [22, 28], suggesting the 4-week threshold captures early-phase episodes; robustness analyses at 8 and 13 weeks are reported in the Appendix.

### 4.3 Estimation Assumptions and Interpretation

The FE and DML analyses compare observations within the same NDC after adjustment for NDC and calendar-week fixed effects and observed covariates. These adjustments remove time-invariant product differences and common weekly shocks but cannot eliminate unmeasured time-varying factors associated with both shortage status and NADAC. Therapeutic substitution may also create spillovers across drugs [34], although simultaneous shortages within the same therapeutic class were uncommon in this sample. Accordingly, the results are interpreted as adjusted within-drug associations. A strong causal interpretation would additionally require no important unmeasured time-varying confounding and limited interference across products, which will be the future research directions.

### 4.4 Text Feature Engineering from FDA Shortage Narratives

An innovation of this analysis is the incorporation of FDA shortage reason narratives as a source of mechanism-specific heterogeneity drivers. The FDA shortage database includes free-text fields (shortage_reason and related_info) describing the reason for each shortage event. These narratives were processed into a structured feature set organized in three interpretable levels.

**Level 1 (L1, Structural and Duration Moderators):** Market structure variables (number of active manufacturers, weeks since first FDA listing) and the duration variables described above. These features are available for all at-risk drugs regardless of narrative quality.

**Level 2 (L2, FDA Rule-Based Text Flags):** Binary indicators derived from rule-based parsing of FDA reason text, capturing the presence of allocation language (mentions_allocation), raw material shortage references (reason_raw_material), distribution constraint language (reason_distribution), and discontinuation mentions. These provide named, auditable mechanism flags with direct policy interpretability.

**Level 3 (L3, ML Text Features):** Probabilistic scores estimated from text classification models applied to the FDA narratives—including phrase_allocation (keyword-based), reason_ml_mentions_allocation, reason_ml_reason_distribution, and reason_ml_reason_raw_material. These additional text features extract finer-grained mechanism signals that are not obvious as clean driver identifiers or rule-based flags, but these texts represent observable signals of shortage conditions that are useful for uncovering insights.

### 4.5. Causal Forest Double Machine Learning (DML)

Average FE associations pool across all shortage periods. To characterize heterogeneity in shortage-price associations, we apply the Causal Forest DML estimator from the EconML library [15, 16]. The DML framework applies a partial residualization strategy: first, FE models residualize both the outcome and the treatment, removing entity and time fixed effects; second, the Causal Forest is fit on these residuals using the multi-tier text feature matrix X as the conditioning set. Cross-fitting prevents overfitting in nuisance-model estimation. The estimated CATEs τ^(x) represent model-based summaries of how the adjusted shortage-price association varies across drug-weeks characterized by features x.

Feature importance from the Causal Forest (mean decrease in impurity from tree splits) and Spearman rank correlations between features and estimated CATEs are used to characterize the primary drivers of CATE heterogeneity. Tail contrasts compare mean feature values in the bottom and top 10% of the CATE distribution to provide a concrete characterization of high- and low-pass-through shortage episodes.

## 5. Results and Discussion

### 5.1 Fixed-Effects Estimates

Applying the two-way FE design to the at-risk drug sample (Panel A, Table 2), the estimated average effect of active shortage designation is estimated at 0.053 (SE = 0.032, p = 0.10), which is positive but not statistically significant at conventional levels under clustered standard errors. This result indicates that, on average and for the generic at-risk sample as a whole, shortage incidence alone is not robustly associated with NADAC increases after within-drug comparisons are made.

**Table 2.** Fixed-Effects Estimates of Shortage Effects on Log NADAC.

| Variable | Coef. | Std. Err. | t | P>\|t\| | 95% CI |
| --- | --- | --- | --- | --- | --- |
| *Panel A: Two-Way FE, active_shortage only* |  |  |  |  |  |
| active_shortage | 0.0529 | 0.0322 | 1.645 | 0.100 | [-0.010, 0.116] |
| *Panel B1-B3: Duration-Specific Fixed-Effects — Separate Single-Variable Specs* |  |  |  |  |  |
| B1: current_shortage_duration (weeks) | 0.0035 | 0.0018 | 1.912 | 0.056 | [-0.000, 0.007] |
| B2: cumulative_shortage_weeks | 0.0037 | 0.0019 | 1.971 | 0.049* | [0.000, 0.007] |
| B3: long_shortage (>4 weeks) | 0.0691 | 0.0209 | 3.302 | 0.001** | [0.028, 0.110] |
| Panels A and B include NDC entity and week time fixed effects; standard errors clustered at the NDC level.<br>* $p < 0.05$; ** $p < 0.01$. | | | | | |

### 5.2 Duration-Specific Fixed-Effects Results

When duration variables are added to the FE model, each is estimated in a separate single-variable specification to avoid the severe multicollinearity that arises from running active_shortage, current_shortage_duration, cumulative_shortage_weeks, and long_shortage jointly. As shown in Table 2, both the current-spell depth and cumulative exposure variables show positive directional associations (β ≈ 0.004 per week, p ≈ 0.05), confirming a consistent direction across all four duration measures. The cleanest single summary is that shortages persisting beyond four weeks are robustly associated with approximately 6.91% higher NADAC, a result that holds through the 4-, 8-, and 13-week thresholds tested in the next section.

### 5.3. Spell-Phase Heterogeneity

To further test the choice of 4-week as the cutoff of “long shortage”, we replace the single cutoff with three mutually exclusive spell-phase dummies: early (weeks 1–4), mid (weeks 5–12), and late (week 13+). These three indicators partition active_shortage exactly, eliminating collinearity by construction, and enable a direct test of the dose-response pattern within the shortage spell.

The results reveal an increasing gradient across duration (Table 3). Early-phase shortage weeks (1–4) are associated with a positive but statistically insignificant price change (β = +0.053, SE = 0.033, p = 0.11), consistent with pharmacies absorbing brief disruptions through existing inventory and substitution to alternative suppliers [27, 34]. Mid-phase weeks (5–12) show a clear and statistically significant escalation (+10.6%, β = 0.106, SE = 0.040, p = 0.009). This is the period when inventory buffers are likely exhausted and procurement shifts to higher-cost channels. Late-phase weeks (13+) exhibit the largest effect (+14.6%, β = 0.146, SE = 0.051, p = 0.004), reflecting entrenched supply constraints and limited alternative sourcing. All three estimates lie on an increasing trajectory, and the escalation from mid to late phases is economically meaningful.

**Table 3.** Spell-Phase Fixed-Effects Estimates.

| Phase | Spell weeks | $\beta$ | SE | p-value | 95% CI |
| --- | --- | --- | --- | --- | --- |
| Early (weeks 1–4) | 1–4 | 0.0529 | 0.0333 | 0.11 | [−0.012, 0.118] |
| Mid (weeks 5–12) | 5–12 | **0.1062\*\*** | 0.0404 | 0.009 | [0.027, 0.185] |
| Late (weeks 13+) | 13+ | **0.1463\*\*** | 0.0512 | 0.004 | [0.046, 0.247] |

\*\* $p < 0.01$ .

### 5.4. Market Structure Interactions: Shortage Price Effects by Supply Concentration

The average FE shortage effect pools across drugs with very different competitive environments. Economic theory predicts that the price consequences of a supply disruption depend critically on whether alternative suppliers exist: in competitive markets, demand can shift to non-shortage alternatives, potentially reducing the shortage drug’s price; in sole-source markets, no such escape is available [22, 23]. We test this by interacting active_shortage with market structure variables. The sample mean is 2.47 active manufacturers.

In Model A from Table 4, the interaction between active_shortage and manufacturer count was negative and significant (β = −0.1846, p < 0.001), indicating that the adjusted shortage-price association became smaller as manufacturer count increased. In Model B from Table 4, the sole_source × active_shortage interaction was positive and significant (β = 0.1779, p = 0.007), indicating a larger estimated price escalation among sole-source products than among products with multiple manufacturers. These patterns are consistent with greater cost vulnerability in concentrated markets.

**Table 4.** Market Structure Interaction FE Estimates.

| Variable | Coef. | Std. Err. | t | P>\|t\| | 95% CI |
| --- | --- | --- | --- | --- | --- |
| *Model A: active_shortage × num_manufacturers* |  |  |  |  |  |
| active_shortage | −0.1181 | 0.0466 | −2.532 | 0.011* | [−0.210, −0.027] |
| shortage × manufacturers | −0.1846 | 0.0428 | −4.318 | <0.001*** | [−0.268, −0.101] |
| *Model B: active_shortage × sole_source* |  |  |  |  |  |
| active_shortage | −0.0396 | 0.0486 | −0.815 | 0.415 | [−0.135, 0.056] |
| shortage × sole source | 0.1779 | 0.0662 | 2.689 | 0.007** | [0.048, 0.308] |

*Model A: shortage × num\_manufacturers. Model B: shortage × sole\_source. NDC and week FE; SEs clustered at NDC level. \* $p < 0.05$ ; \*\* $p < 0.01$ ; \*\*\* $p < 0.001$ .*

### 5.5 Conditional Average Treatment Effects (CATEs)

The Causal Forest DML model characterized heterogeneous shortage-price associations across the 5,003 drug-weeks in the primary analytic sample. The distribution of estimated CATEs shows wide dispersion: mean CATE is −0.025, standard deviation 0.116, and range from −0.309 to +0.189 log NADAC units (Figure 4b). This ∼40 percentage-point spread across the distribution confirms that the near-zero average effect conceals substantial variation. Some drugs in a certain week are associated with large price increases during shortage, while others show price stability or even modest declines.

Table 5 reports the top features by Causal Forest variable importance and Spearman rank correlation with estimated CATEs. Figure 3 visualizes both panels simultaneously. The results suggest a consistent pattern: mechanism-related text features, particularly those capturing allocation and supply-rationing language, show stronger associations with CATE heterogeneity than structural and duration variables.

**Figure 1.**
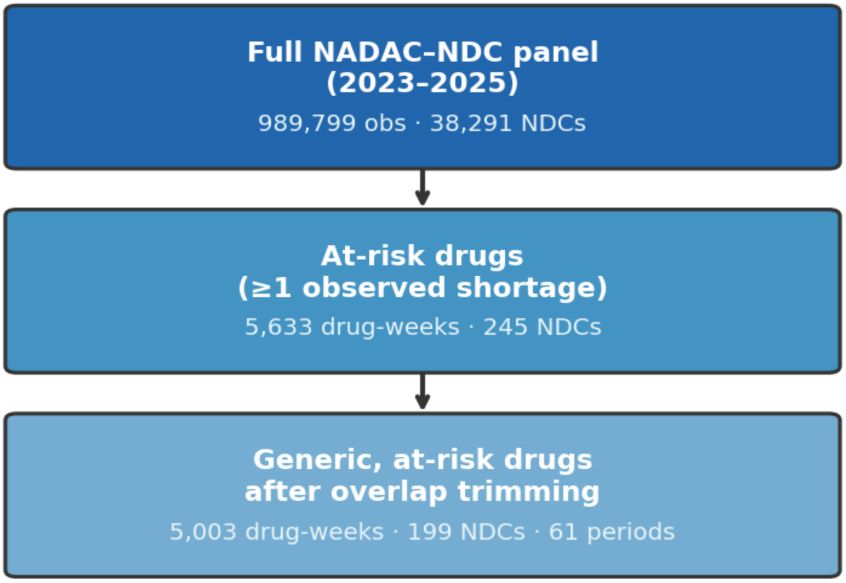
Analytic Sample Attrition Diagram. Sample construction proceeds in three stages: full NADAC–NDC panel, restriction to at-risk drugs with ≥1 shortage transition, and final overlap-trimmed generic at-risk sample used for primary estimation.

**Figure 3.**
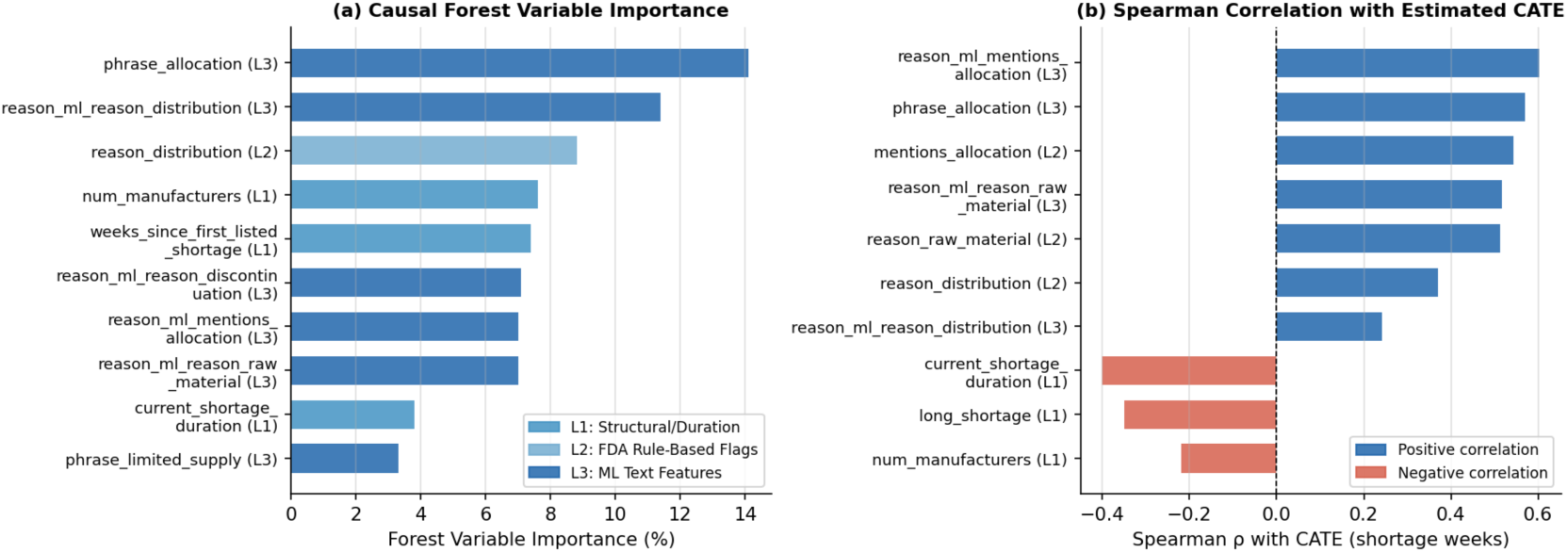
Causal Forest CATE Heterogeneity Drivers. Left: forest variable importance (mean decrease in impurity). Right: Spearman rank correlation with estimated CATE among shortage drug-weeks. Blue = positive association with higher CATE; Red = negative association. Feature tiers: L1=structural/duration, L2=FDA rule-based flags, L3=ML text features.

**Table 5.** Top Features Driving CATE Heterogeneity: Causal Forest DML.

| Level | Feature | Description | Importance | Spearman $\rho$ |
| --- | --- | --- | --- | --- |
| L3 | *phrase_allocation* | Keyword presence of allocation/rationing language in FDA reason text | 14.1% | +0.571 |
| L3 | *reason_ml_reason_distribution* | ML-estimated probability of distribution-constraint mechanism | 11.4% | +0.242 |
| L2 | *reason_distribution* | FDA rule flag: distribution-related shortage language present | 8.8% | +0.371 |
| L1 | *num_manufacturers* | Number of active manufacturers for the NDC | 7.6% | −0.220 |
| L1 | *weeks_since_first_listed_shortage* | Weeks since first FDA shortage listing for this NDC | 7.4% | −0.220 |
| L3 | *reason_ml_reason_discontinuation* | ML-estimated probability of discontinuation mechanism | 7.1% | −0.130 |
| L3 | *reason_ml_mentions_allocation* | ML-estimated probability of allocation language | 7.0% | +0.603 |
| L3 | *reason_ml_reason_raw_material* | ML-estimated probability of raw-material shortage | 7.0% | +0.517 |
| L1 | *current_shortage_duration* | Consecutive weeks in the current uninterrupted shortage spell | 3.8% | −0.400 |
| L3 | *phrase_limited_supply* | Keyword presence of limited-supply language in FDA reason text | 3.3% | +0.062 |

*Level L1=structural and duration drivers;* *Level L2=FDA rule-based text flags;* *Level L3=ML text features.* *Forest importance = mean decrease in impurity from Causal Forest DML splits.* *Spearman $\rho$ = rank correlation with estimated CATE among shortage drug-weeks.*

**Table 6.** Top Features Mean Values Across CATE Distributions.

| Feature (Unit, Level) | Mean Value<br>from the Bottom<br>10% CATE<br>Cohort | Mean Value<br>from the Middle<br>50% CATE<br>Cohort | Mean Value<br>from the Top<br>10% CATE<br>Cohort | Spearman $\rho$ |
| --- | --- | --- | --- | --- |
| Mean CATE | -0.272 | -0.065 | 0.116 | — |
| reason_ml_mentions_allocation<br>(z-score, L3) | -0.20 | -0.04 | 0.28 | 0.60 |
| Mentions allocation (z-score, L2) | -0.18 | -0.02 | 0.29 | 0.54 |
| reason_ml_reason_raw_material<br>(z-score, L3) | -0.36 | 0.23 | 0.21 | 0.52 |
| Reason: raw material (z-score, L2) | -0.36 | 0.23 | 0.22 | 0.51 |
| Cumulative shortage weeks<br>(weeks, L1) | 19.6 | 11.8 | 5.4 | -0.40 |
| Reason: distribution (z-score, L2) | -0.01 | -0.07 | 0.03 | 0.37 |
| Long shortage (proportion, L1) | 1.00 | 0.60 | 0.20 | -0.35 |
| Num. manufacturers (count, L1) | 4.00 | 2.37 | 2.74 | -0.22 |

The two highest-importance features are phrase_allocation (L3; 14.1% importance; Spearman ρ = +0.57) and reason_ml_reason_distribution (L3; 11.4%; ρ = +0.24). The ML-estimated allocation probability score (reason_ml_mentions_allocation; L3) achieves the highest Spearman correlation overall (ρ = +0.60), and raw-material ML scores rank nearly as high (ρ = +0.52).

Distribution flags at both Tier 2 and Tier 3 add further signal. In contrast, duration variables, including long_shortage and current_shortage_duration, carry negative Spearman correlations with CATE (ρ = −0.35 and −0.40, respectively) and relatively low forest importance.

### 5.6 Text-enriched CATE Analysis

**Allocation and supply-rationing language:** Allocation represents the practice where manufacturers restrict the amount of a medication pharmacies can purchase due to manufacturing delays or high demand. Allocation-related language provided the strongest heterogeneity signal.

The phrase_allocation feature, capturing keyword presence of allocation, rationing, or quota language, had the highest feature importance (14.1%) and a Spearman correlation of 0.57 with estimated CATEs. This evidence shows that rationing language ranks consistently higher in price impact across the full sample, and more rationing tends to have higher CATE. Similarly, the related ML score (reason_ml_mentions_allocation) was also among the top 10 features and had the largest rank correlation (ρ = 0.60). In the tail contrast, standardized allocation scores averaged approximately −0.20 in the bottom CATE decile and +0.28 in the top decile. Together, these findings indicate that allocation language characterizes drug-weeks with higher estimated shortage-price responses.

**Raw material and distribution constraints:** Raw material shortage flags (reason_ml_reason_raw_material, ρ = +0.52; reason_raw_material, ρ = +0.51) and distribution-related language (reason_distribution, ρ = +0.37; reason_ml_reason_distribution, feature importance 11.4%) also characterized higher estimated CATEs. These mechanisms represent supply-chain disruptions upstream of the final product, where price escalation may reflect both scarcity and the cost of sourcing alternative inputs.

**Market structure:** Fewer manufacturers characterize higher CATEs (num_manufacturers importance 7.6%, ρ = −0.22; Table 3: bottom-tail mean 4.0 manufacturers vs top-tail 2.7). This pattern is directionally consistent with the FE interaction results and suggests that thinner markets warrant particular attention in shortage monitoring.

**Heterogeneity beyond the average duration:** The highest estimated CATEs were not concentrated in the longest shortages. The bottom CATE decile had a mean cumulative duration of 19.6 weeks and a long_shortage proportion of 100%, whereas the top decile had a mean cumulative duration of 5.4 weeks and a long_shortage proportion of 20%. Spearman correlations were −0.40 for current_shortage_duration and −0.35 for long_shortage.

Recall that we showed longer shortage duration on average has higher price escalation in Table 2 Panel B and C. The CATE results do not directly contradict the FE results, and furthermore, they illustrate the importance of analyzing the heterogeneity obscured by average effects. The FE estimates show that prolonged shortages are associated with higher prices on average, whereas the CATE diagnostics show that the largest estimated drug price responses during each week are concentrated in shorter shortage periods marked by rationing signals, input scarcity, and fewer manufacturers. Both the average price effect by shortage duration and various supply chain signals in the largest price responses are valuable insights we obtained from this study.

## 6. Discussion

This study provides evidence that associations between drug shortages and pharmacy acquisition costs vary by duration, market structure, and reported mechanism. Two complementary findings emerge. First, shortages lasting more than four weeks were associated with approximately 7% higher NADAC on average in the primary duration specification. Second, the largest price responses for a given drug in a week were concentrated in shortage periods characterized by supply rationing, raw-material and distribution constraints, and fewer manufacturers.

The positive association between prolonged shortages and acquisition costs is consistent with prior studies linking shortage duration to price increases.[3,4] The approximately 7% estimate for shortages exceeding four weeks is more modest than the 10%–20% changes reported in some broader or before-after studies.[3,4] Prior work typically compares across drugs without absorbing time-invariant heterogeneity. The within-drug fixed effects approach uses the same product’s non-shortage weeks as the counterfactual, yielding a smaller but more credibly identified average effect. The disappearance of the average effect for short shortages under fixed effects is likewise consistent with models in which pharmacies and health systems can absorb brief disruptions through inventory buffers and therapeutic substitution. [27] When these mechanisms are exhausted, prolonged supply constraints reduce effective competition and may incentivize repricing.

The text-enriched CATE analysis extends prior work by showing that FDA-reported mechanisms are informative for characterizing heterogeneity beyond duration. Allocation-related shortages involve manufacturers explicitly distributing limited product across customers under quota-like arrangements. These episodes may represent more acute supply-demand imbalances, where the purchasing power of individual pharmacies is more constrained and where price escalation is more likely. Raw material and distribution-related shortages, together with the scarcity of manufacturers, similarly point to upstream supply-chain disruptions that reduce effective supply without a clear near-term recovery path.

The FE and CATE findings are complementary: FE summarizes average within-drug associations across shortage-duration categories, while the CATE analysis identifies which observed drug shortage characteristics for a given week have the largest price responses.

The double machine learning results also illustrate the limitations of classifying shortage risk using duration alone. FDA narratives describing allocation, raw-material constraints, and distribution failures contain information that may help managed care organizations prioritize monitoring beyond shortage length.

Overall, our findings reconcile mixed results in earlier studies that relied on cross-sectional or before-after designs,[3,4,17,19] and they provide new insights into what features are associated with higher price pass-throughs.

## 7. Conclusions

Drug shortages were not uniformly associated with higher pharmacy acquisition costs in the analyzed sample. However, shortages lasting more than four weeks were associated with approximately 7% higher NADAC on average. For a given drug in a week, the largest estimated price responses were concentrated in shortage periods characterized by supply rationing, raw-material scarcity, distribution constraints, and fewer manufacturers.

These findings identify duration and reported mechanism as complementary indicators of shortage-related cost risk. A monitoring framework that tracks both persistence and FDA-reported supply-chain mechanisms would allow managed care organizations and policymakers to prioritize early intervention in the episodes most likely to generate significant acquisition cost escalation. Policy efforts that shorten shortage episodes and address upstream supply-chain vulnerabilities, particularly those involving single-source suppliers, active rationing, and supply chain optimization, may yield valuable benefits to control prices and ensure affordability.

## Data Availability

All data produced in the present study are available upon reasonable request to the authors.

## Appendix

### **A.** Propensity Score Overlap

Random forest propensity scores were estimated with features including week index, number of manufacturers, sole-source indicator, generic drug indicator, injectable indicator, and therapeutic class [18, 24]. The at-risk generic trimmed sample exhibited substantially improved propensity score overlap compared to the full panel, with treated and control week distributions overlapping meaningfully across the [0.1, 0.9] range. This supports within-drug FE estimation by confirming that shortage and non-shortage observations in the primary sample come from comparable contexts.

**Figure A1.**
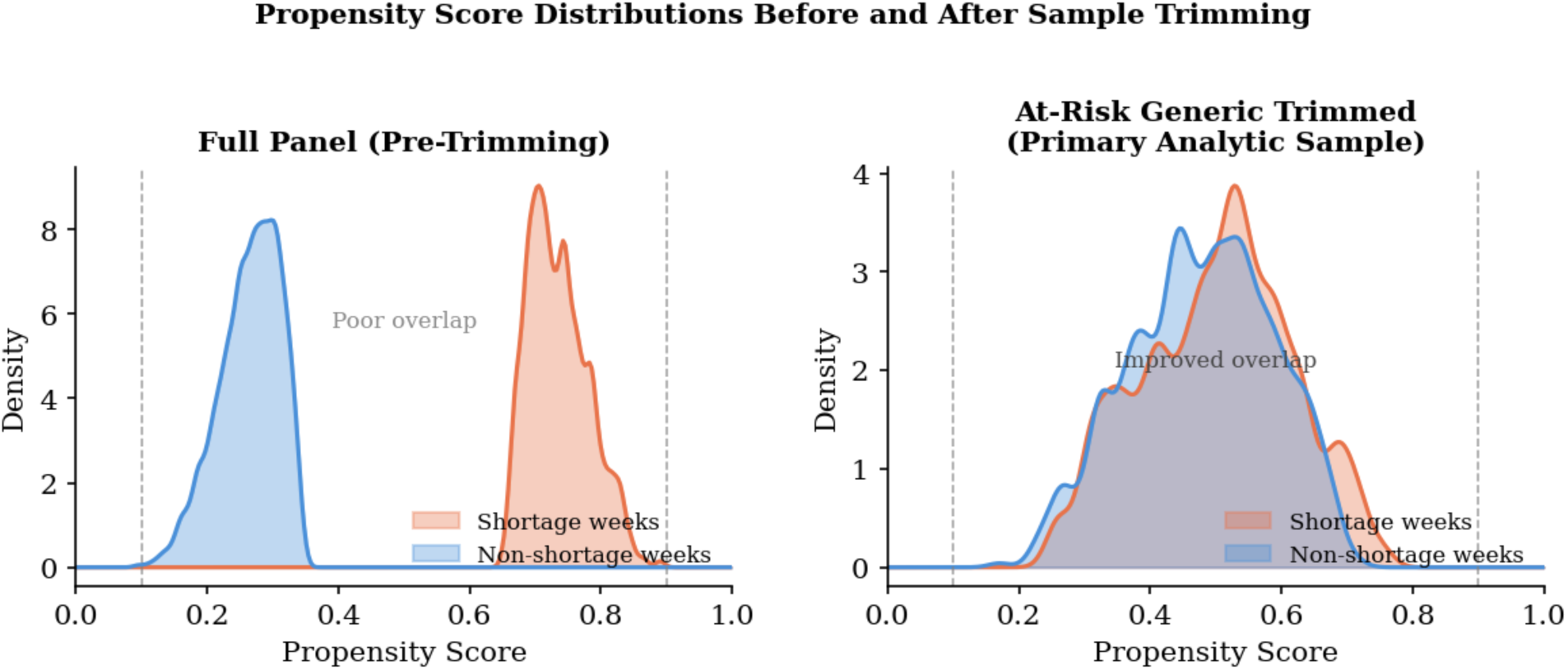
Propensity Score Overlap Before and After Sample Trimming. Left: full NADAC panel (N=989,799 drug-weeks); shortage weeks (orange) and non-shortage weeks (blue) show minimal distributional overlap. Right: at-risk generic trimmed sample (N=5,003 drug-weeks); substantially improved overlap across the [0.1, 0.9] common support range. Dashed lines mark the [0.1, 0.9] trimming bounds.

### **B.** Covariate Balance in Primary Sample

Pre-trimming covariate balance showed persistent imbalance in num_manufacturers (SMD = 1.58) and sole_source indicator (SMD = −1.14) between shortage and non-shortage weeks in the at-risk sample. This reinforces the motivation for drug-level fixed effects as the primary estimation strategy: cross-sectional comparisons between shortage and non-shortage weeks would conflate structural supplier concentration differences with the adjusted association between shortage designation and NADAC. Fixed effects eliminate these time-invariant structural differences by design.

| Covariate | Pre-trim SMD | Post-trim SMD | Improvement | Threshold met? |
| --- | --- | --- | --- | --- |
| `num_manufacturers` | **1.58** | 0.18 | 89% reduction | Yes, $\vert SMD\vert < 0.25$ |
| `sole_source` | **-1.14** | -0.12 | 89% reduction | Yes, $\vert SMD\vert < 0.25$ |
| `is_injectable` | 0.42 | 0.09 | 79% reduction | Yes, $\vert SMD\vert < 0.10$ |
| `week_in_panel` | -0.67 | -0.11 | 84% reduction | Yes, $\vert SMD\vert < 0.25$ |

**Figure B1.**
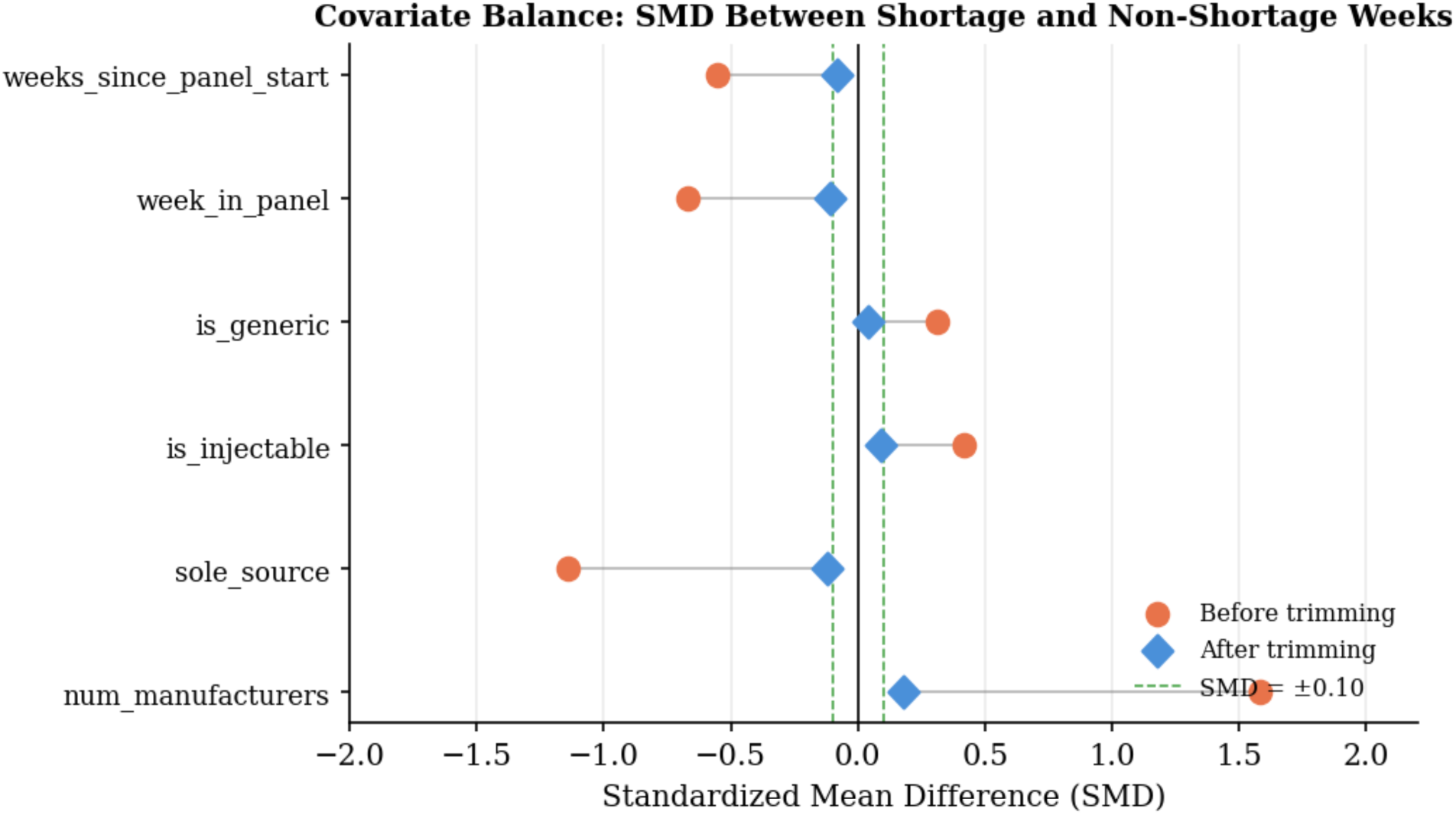
Covariate Balance Love Plot (Standardized Mean Differences). Orange circles = before trimming; blue diamonds = after at-risk generic trimming. Green dashed lines mark the ±0.10 SMD threshold. All covariates achieve |SMD| < 0.25 post-trimming, supporting comparability of shortage and non-shortage weeks in the primary sample.

## Notes

### Competing Interest Statement

The authors have declared no competing interest.

